# ONE ASSAY TO TEST THEM ALL: COMPARING MULTIPLEX ASSAYS FOR EXPANSION OF RESPIRATORY VIRUS SURVEILLANCE

**DOI:** 10.1101/2023.01.19.23284806

**Authors:** Narjis Boukli, Claude Flamand, Kim Lay Chea, Leangyi Heng, Seangmai Keo, Kimhoung Sour, Sophea In, Panha Chhim, Bunthea Chhor, Lomor Kruy, Jelena D M Feenstra, Manoj Gandhi, Obiageli Okafor, Camilla Ulekliev, Heidi Auerswald, Viseth Srey Horm, Erik A Karlsson

## Abstract

**Background:** Molecular multiplex assays (MPAs) for simultaneous detection of severe acute respiratory syndrome coronavirus 2 (SARS-CoV-2), influenza and respiratory syncytial virus (RSV) in a single RT-PCR reaction reduce time and increase efficiency to identify multiple pathogens with overlapping clinical presentation but different treatments or public health implications.

**Methods:** Clinical performance of XpertXpress^®^ SARS-CoV-2/Flu/RSV (Cepheid, GX), TaqPath™ COVID-19, FluA/B, RSV Combo kit (Thermo Fisher Scientific, TP), and PowerChek™ SARS-CoV-2/Influenza A&B/RSV Multiplex RT-PCR kit II (KogeneBiotech, PC) was compared to individual Standards of Care (SoC). Thirteen isolates of SARS-CoV-2, human seasonal influenza, and avian influenza served to assess limit of detection (LoD). Then, positive and negative residual nasopharyngeal specimens, collected under public health surveillance and pandemic response served for evaluation. Subsequently, comparison of effectiveness was assessed.

**Results:** The three MPAs confidently detect all lineages of SARS-CoV-2 and influenza viruses. MPA-LoDs vary from 1-2 Log10 differences from SoC depending on assay and strain. Clinical evaluation resulted in overall agreement between 97% and 100%, demonstrating a high accuracy to detect all targets. Existing differences in costs, testing burden and implementation constraints influence the choice in primary or community settings.

**Conclusion:** TP, PC and GX, reliably detect SARS-CoV-2, influenza and RSV simultaneously, with reduced time-to-results and simplified workflows. MPAs have the potential to enhancediagnostics, surveillance system, and epidemic response to drive policy on prevention and control of viral respiratory infections.

**IMPORTANCE:** Viral respiratory infections represent a major burden globally, weighed down by the COVID-19 pandemic, and threatened by spillover of novel zoonotic influenza viruses. Since respiratory infections share clinical presentations, identification of the causing agent for patient care and public health measures requires laboratory testing for several pathogens, including potential zoonotic spillovers. Simultaneous detection of SARS-CoV-2, influenza, and RSV in a single RT-PCR accelerates time from sampling to diagnosis, preserve consumables, and streamline human resources to respond to other endemic or emerging pathogens. Multiplex assays have the potential to sustain and even expand surveillance systems, can utilize capacity/capability developed during the COVID-19 pandemic worldwide, thereby strengthening epidemic/pandemic preparedness, prevention, and response.

## BACKGROUND

Aside from novel coronavirus disease 2019 (COVID-19), respiratory infections with viral pathogens remain a major global burden [1–3]. Since numerous respiratory viruses circulate concurrently with similar clinical presentations, diagnosis requires laboratory testing for several pathogens. Any delays in accurate and timely identification can compromise patient care [4].

Real-time polymerase chain reaction (RT-PCR) on upper respiratory tract (URT) swabs is the gold standard for diagnosis of viral respiratory infections (VRIs) [5]. Between 2020 and 2022, public health measures to constrain COVID-19 significantly altered incidence of VRIs [6]. However, with reduction of restrictions and fatigue over prevention behaviors, both influenza and respiratory syncytial virus (RSV) are resurging [7,8]. Co-infections can increase severity and mortality [9,10]. In addition, spillovers of novel zoonotic influenza viruses continually represent a human threat [11,12]. Funding issues, disruptions in reagent procurement and supply chains, and inadequate human resources reduce diagnostic testing capacity, especially under pandemic conditions. Therefore, improvement of VRI surveillance needs to account not only for multiple pathogens and their potential genetic and seasonal changes, but also for human resources, capacity, and cost.

Molecular multiplex assays (MPAs) allowing detection of several pathogens in a single RT-PCR have demonstrated utility for diagnostics of influenza and RSV [13]. Early in the COVID-19 pandemic, manufacturers modified existing MPAs to simultaneously detect severe acute respiratory syndrome coronavirus-2 (SARS-CoV-2) [5,14]. Both the United States and Wales recommend MPA integration to detect SARS-CoV-2 and influenza in their public health strategies [15,16]. Considering the co-circulation of respiratory viruses and suggested expansion of testing in global surveillance, MPAs may be an attractive option [17,18]. However, viral evolution, genetic bottlenecks, and emergence of novel avian influenza (AIV) strains could impair viral detection [19,20].

Comparison between MPAs and standard protocols allows evaluation of the clinical performance, as well as cost and testing burden for three commercial multiplex RT-PCR assays intended to simultaneously detect SARS-CoV-2, influenza, and RSV.

## METHODS

### Assays

Three MPAs available and easily implemented in Cambodia were performed according to manufacturers’ protocols (Table 1).

**TABLE 1:**
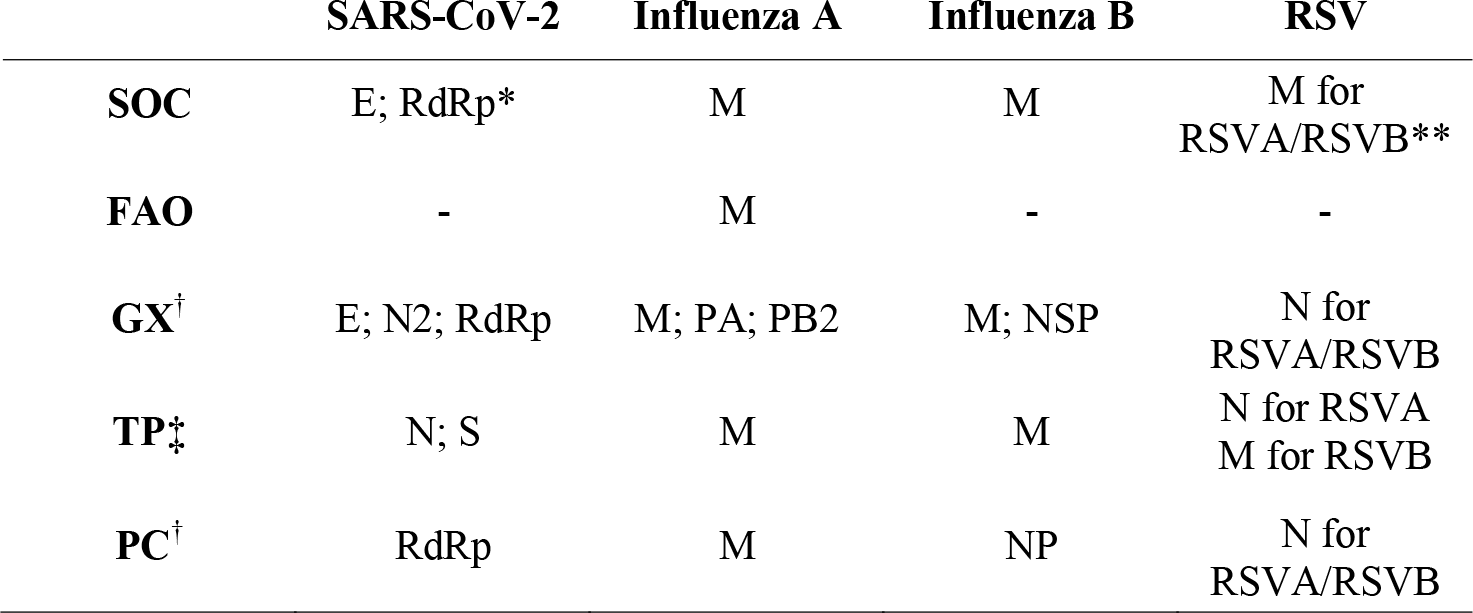
Genes targeted for each virus and each assay SARS-CoV-2: severe acute respiratory syndrome coronavirus 2; IAV: Influenza A virus ; IBV: Influenza B virus ; RSVA/B: respiratory syncytial virus A/B; SoC: standard of care; FAO: Food and Agriculture Organization of United Nations; GX: Xpert^®^ Xpress SARS-CoV-2, Flu, RSV Kit; TP: TaqPath™ COVID-19, FluA/B, RSV Combo Kit; PC: PowerChek™ SARS-CoV-2, Influenza A&B, RSV Multiplex Realtime PCR Kit II; E: Envelop; M: Matrix; N: nucleocapsid; NP: nucleoprotein; NSP: non-structural protein; PA: polymerase acidic protein; PB2: polymerase basic protein; RdRp: RNA-dependent RNA polymerase; S: spike. MPAs target one, two or three genes for detection of each SARS-CoV-2, IAV, IBV and RSV. * SoC use two sets of primers and probes performed in two separate wells/PCR runs for each sample for detection of SARS-CoV-2; ** SoC use different optical channels to detect the RSV targets and then provide separate results for RSVA and RSVB; † GX and PC use separate optical channels to detect SARS-CoV-2, IAV, IBV and RSV and provide results for each virus separately; ‡ TP uses one optical detection channel for the detection of IAV and IBV and provides a combined result for influenza A/B, and similarly one optical channel is used for detection of RSVA and RSVB providing a combined result for RSV.

1. **XpertXpress**^**TM**^ **SARS-CoV-2/Flu/RSV test** (GX) (Cepheid, CA, USA), a closed unitary MPA, integrates specimen extraction, RT-PCR, and target detection [21]. A GeneXpert Xpress XVI-16 instrument (Cepheid) served to run cartridges, and instrument software generated result interpretation.
2. **TaqPath**^**TM**^ **COVID-19, FluA/B, RSV Combo Kit** (TP) (Thermo Fisher Scientific, MA, United States) is a MPA with two targets for each virus [22]. RT-PCR was performed on the QuantStudio™ 5 RT-PCR Instrument, 0.2 mL block (Applied Biosystems, MA, USA) and results were analyzed using the Pathogen Interpretive Software CE-IVD Edition v1.1.0 (Applied Biosystems).
3. **PowerChek**^**TM**^ **SARS-CoV-2, Influenza A&B, RSV Multiplex Real-time PCR Kit II** (PC) (KogeneBiotech, Inchon, Korea), a MPA with one targeted gene for each virus [23], was performed on the CFX96™ RT-PCR Detection System (Bio-Rad Laboratories, CA, USA) and results analyzed with CFX96™ software.

Standard of care assays (SoC) utilized at IPC for the detection of SARS-CoV-2 (CoV-SoC), influenza A virus (IAV-SoC), influenza B virus (IBV-SoC) and RSV (RSV-SoC), consisting of single RT-PCR tests (Table 1), served as reference [24–27]. In addition, IAV samples were tested using Food and Agriculture Organization of United Nations (FAO) recommended primers and probes developed by the Australian Center for Disease Prevention for the detection of M gene from avian influenza viruses (AIV) in Asia [28]. All SoC and FAO were performed on a CFX96™ instrument and results analyzed with the corresponding software.

### Study specimens

In-house Cambodian viral isolates, including several variants of SARS-CoV-2 and subtypes of human seasonal influenza, and AIV (Table 2) were heat-inactivated and used to assess the limit of detection (LoD) of each assay. For each isolate, a serial-dilution was prepared in standard Viral Transport Media (VTM) and stored at -70°C. Immediately after thawing, 300µl of sample was tested with GX and 400µl was extracted with the MagMAX™ Viral/Pathogen II Nucleic Acid Isolation Kit on a KingFisher Flex system (Thermo Fisher Scientific), using the volume recommended by TP instructions for use, and RNA eluted with 50µl nuclease-free water. Each 10-fold dilution was tested in triplicate with SoC. End-point dilution was defined as lowest dilution at which all replicates were positive. Subsequently, each viral isolate was tested with GX, TP, PC and SoC in parallel on the same day, at the previously determined end-point dilution and a minimum of two half-log_10_ dilutions on either side of the LoD.

**TABLE 2:**
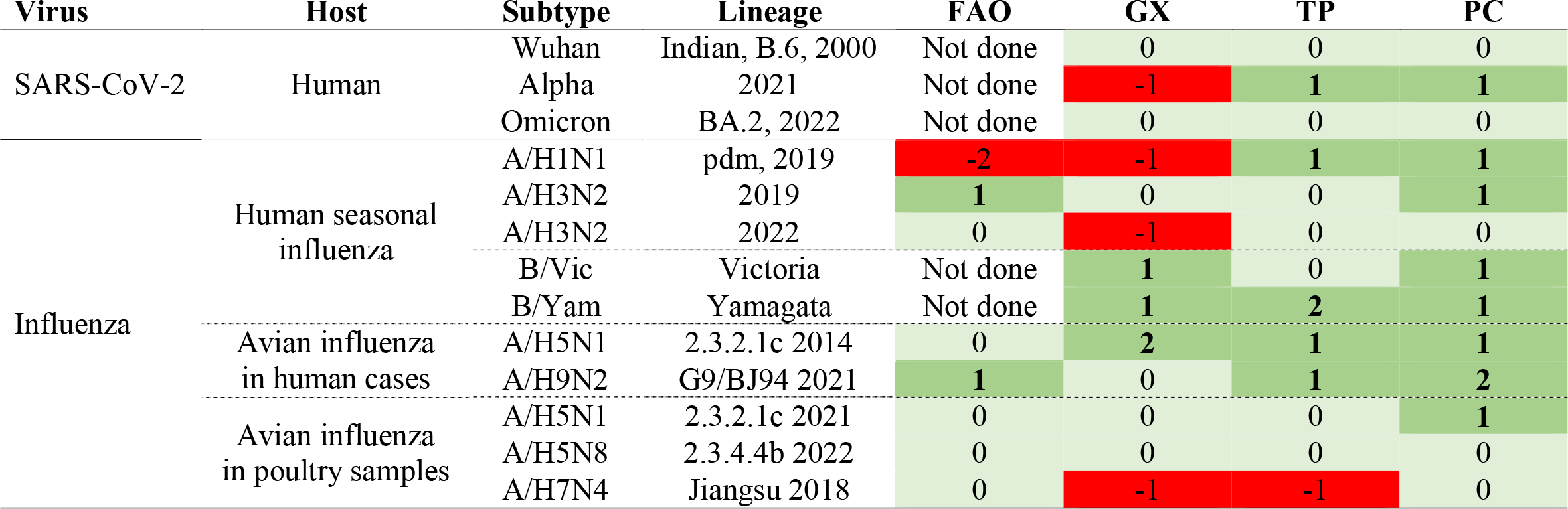
Comparison of limit of detection between evaluated and standard assays FAO: Food and Agriculture Organization of United Nations recommended primers and probes developed by the Australian Center for Disease Prevention for the detection of M gene from avian influenza; GX: Xpert^®^ Xpress SARS-CoV-2, Flu, RSV Kit; TP: TaqPath™ COVID-19, FluA/B, RSV Combo Kit; PC PowerChek™ SARS-CoV-2, Influenza A&B, RSV Multiplex Real-time PCR Kit II; SARS-CoV-2: severe acute respiratory syndrome coronavirus 2. The table presents the difference between SoC and evaluated assay in term of Log10 dilution. Delta LoD resulted in 0 if MPA and SoC had the same LoD (in light green), ≥ 1 if LoD of MPA was lower than SoC (in dark green) and <0 if LoD of MPA was higher than SoC (in red).

To assess clinical accuracy, residual URT specimens collected in VTM were selected based on routine results obtained under public health surveillance for influenza (IAV n= 84, IBV n= 5) and RSV (n=32), and pandemic response for SARS-CoV-2 (n= 58), upon availability and volume of stored samples (supplementary table 1). Different lineages were selected based on molecular and sequencing results. Samples previously tested negative for all targets were also included (n=126). Similar to viral isolates, 300µl of sample were used for GX testing and 400µl for extraction. Extracted RNA served for side-by-side testing with TP, PC, and SoC, performed on the same day. As amount of RNA for each sample was limited to re-test with SoC, routine negative results were utilized for comparison in the following cases: for IAV, IBV, and RSV among the SARS-CoV-2 samples; for IBV and RSV for IAV samples; for IAV and RSV for IBV samples; for SARS-CoV-2 and IAV/IBV among negative samples. Influenza and RSV samples collected during influenza/RSV seasons in 2016-2019 were negative for SARS-CoV-2. However, if one targeted virus was detected with any MPA, the related SoC was performed using the same RNA. For 31/84 IAV specimens, remaining volume was not sufficient to perform GX testing in addition to extraction.

### Statistical analysis

For each assay, individual cycle threshold (Ct) values (Ct-values) and interpretation as positive or negative according to test cut-off were recorded for each viral isolate and clinical sample. Three (SARS-CoV-2; influenza; RSV) results for TP or 4 (SARS-CoV-2; IAV; IBV; RSV) for GX, PC and SoC were provided for each sample. Comparison was performed for each virus individually. Difference between LoD with SoC and each MPA (D-LoD) was calculated for each viral isolate. D-LoD resulted in 0 when MPA and SoC had the same LoD, _≥_ 1 if MPA LoD was higher than SoC and < 0 if MPA LoD was lower than SoC. Sensitivity, specificity, positive and negative predictive values (PPA/NPA) were calculated using STATA statistical software (v12.1, College Station, TX, USA). Overall accuracy to detect viruses in clinical samples for GX, TP and PC was assessed by percent agreement, corresponding to the proportion of identical results between each MPA evaluated and SoC for each virus, and 95% confidence intervals (95% CI).

### Assessment of utility

Total turnaround time per specimen, including extraction, RT-PCR, and interpretation of results were compared. Cost comparison accounted for reagents and shipments to Cambodia at current pricing structures. Other criteria to help drive choice for suitability included the volume of sample for extraction/assay, amount of RNA for RT-PCR, equipment requirements, practicability of interpretation software, result type obtained for each targeted virus.

## RESULTS

### Limit of Detection

The three MPAs consistently detected all selected viral strains with D-LoDs ranging from -2 to +2 Log10 dilutions according to strains and assays (Table 2). A higher D-LoD occurred on GX for 5/13 isolates: SARS-CoV-2 Alpha and Omicron variants and recent A(H1N1), A(H3N2-2022) IAV and A(H9N2) AIV from human sample, but LoD was equivalent or lower for other isolates. TP had equivalent (6/13 isolates) or slightly better LoD (5/13 isolates) compared to SoC except for A(H7N4). For PC, all LoDs were equivalent or slightly better than SoC.

### Performance on clinical samples

Median and range of Ct-values on GX were equivalent to SoC, but lower using TP and PC (Figure 1). TP and PC adequately detected all selected positive samples from all lineages and all negative samples with sensitivity and specificity over 95% (Supplemental Table 2). GX identified all but four samples, for which the test failed to detect RSV. Discordant results occurred in 14 samples on the remaining targets (Supplemental Table 3). Among two samples with RSV/SARS-CoV-2 co-infection, none of the MPAs detected SARS-CoV-2 in the first, and PC failed to detect SARS-CoV-2 in the second sample. Among the 4/32 RSV samples (12.5%) not detected with GX, three were mono-infections. The last one had an IAV/RSV co-infection. TP also failed to detect RSV in this sample. Eight additional samples had a positive result for one target but were not detected by SoC or other MPA: two had positive result only with TP (1 influenza; 1 RSV) and six only with PC (1 SARS-CoV-2; 1 IAV; 3 IBV; 1 RSV).

**FIGURE 1:**
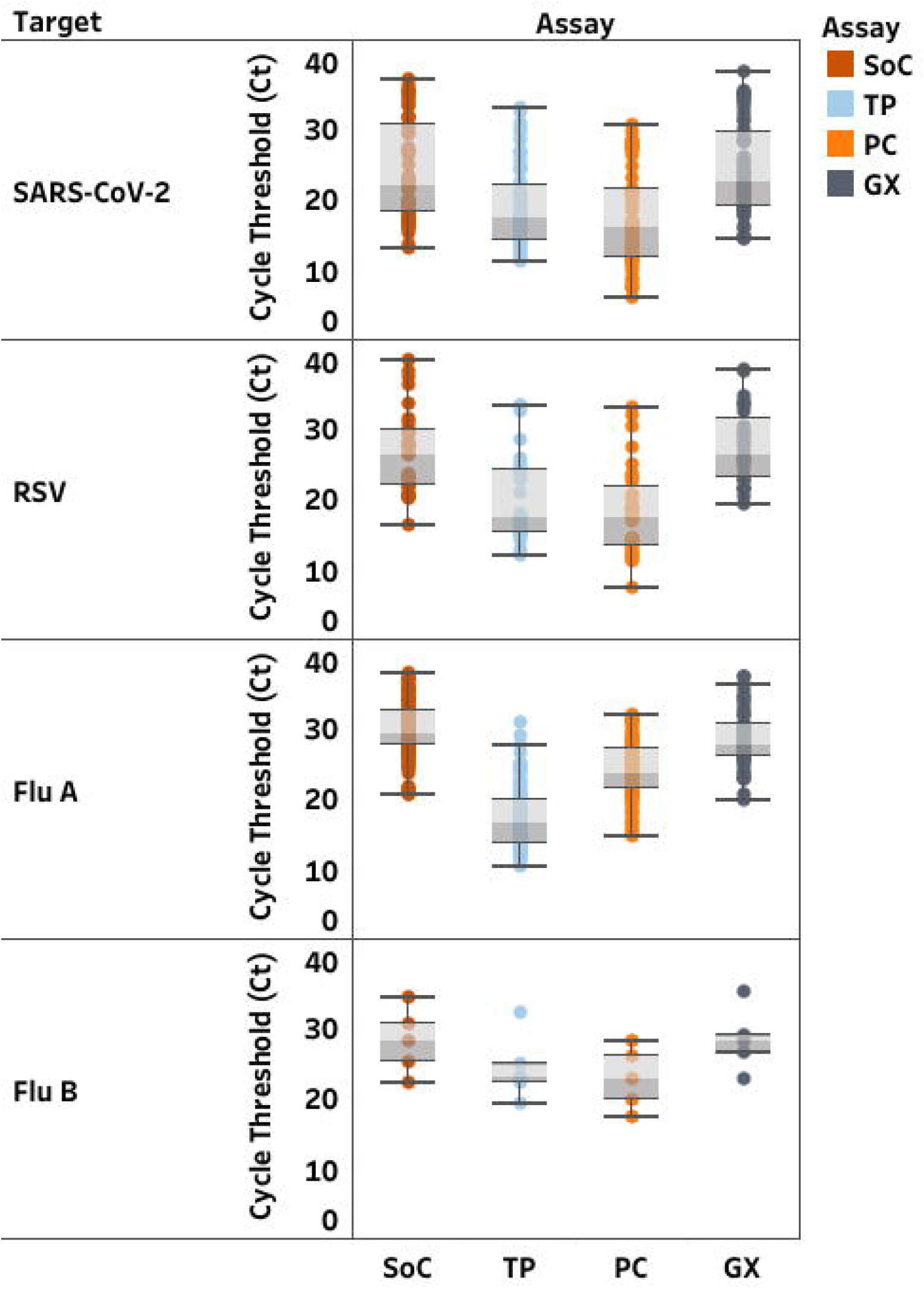
Distribution of cycle threshold (Ct)-values (median; min-max) in clinical samples according to each RT-PCR assay. Standards of Care (SoC) are displayed in dark orange, Thermofisher TaqPath™ COVID-19, FluA/B, RSV Combo Kit (TP) in light blue, Kogene PowerChek™ SARS-CoV-2, Influenza A&B, RSV Multiplex Real-time PCR Kit II (PC) in light orange, Cephied Xpert® Xpress SARS-CoV-2, Flu, RSV Kit (GX) in grey. Ct values are displayed for severe acute respiratory syndrome coronavirus 2 (SARS-CoV-2), respiratory

Overall, positive and negative predictive values (PPV, NPV) ranged between 97% and 100%, except for detection of IBV using PC which dropped to 62.5%. However, overall accuracy between SoC and MPA ranged between 97% and 100% of agreement (Table 3).

**TABLE 3:** Comparison of evaluated assay and standard WHO/GIRS assays currently used in the laboratory SARS-CoV-2: severe acute respiratory syndrome coronavirus 2; GX: Xpert^®^ Xpress SARS-CoV-2, Flu, RSV Kit; TP: TaqPath™ COVID-19, FluA/B, RSV Combo Kit; PC: PowerChek™ SARS-CoV-2, Influenza A&B, RSV Multiplex Real-time PCR Kit II; FAO: Food and Agriculture Organization of United Nations recommended primers and probes developed by the Australian Center for Disease Prevention for the detection of M gene from avian influenza. * TP provides a combine influenza result for IAV and IBV as targets are combined in the same optical detection channel. Therefore, IAV and IBV results were combined for statistical tests.

|  |  | Sensitivity | Specificity | Positive predictive value | Negative predictive value | Overall percent agreement |
| --- | --- | --- | --- | --- | --- | --- |
| SARS-CoV-2 | GX | 96.6 (88.3-99.6) | 100.0 (93.7-100.0) | 100.0 (97.8-100.0) | 98.8 (95.7-99.9) | 99.1 (97.0-99.9) |
|  | TP | 96.6 (88.3-99.6) | 100.0 (98.0-100.0) | 100.0 (93.7-100.0) | 98.9 (96.0-99.9) | 99.2 (97.0-99.9) |
|  | PC | 95.0 (85.9-98.9) | 99.4 (96.9-100.0) | 98.2 (90.6-100.0) | 98.4 (95.3-99.7) | 98.3 (95.8-99.5) |
| Influenza A virus | GX | 100.0 (93.3-100.0) | 100.0 (98.0-100.0) | 100 (93.3-100.0) | 100.0 (98.0-100.0) | 100.0 (98.4-100.0) |
|  | PC | 100.0 (95.7-100.0) | 99.4 (96.9-100.0) | 98.8 (93.6-100.0) | 100.0 (98.1-100.0) | 99.6 (98.0-100.0) |
| Influenza B virus | GX | 100.0 (47.8-100.0) | 100 (98.2-100.0) | 100.0 (47.8-100.0) | 100.0 (98.2-100.0) | 100.0 (98.2-100.0) |
|  | PC | 100.0 (47.8-100.0) | 98.6 (96.1-99.7) | 62.5 (24.5-91.5) | 100.0 (98.3-100.0) | 98.7 (96.2-99.7) |
| InfluenzaA/B | TP* | 100.0 (96.0-100.0) | 100.0 (98.0-100.0) | 100.0 (97.9-100.0) | 100.0 (97.9-100.0) | 100.0 (98.6-100.0) |
| Respiratory syncytial virus | GX | 87.5 (71.0-96.5) | 100.0 (97.8-100.0) | 100.0 (87.7-100.0) | 97.6 (94.0-99.3) | 97.9 (94.8-99.4) |
|  | TP | 96.9 (83.8-99.9) | 100.0 (97.9-100.0) | 100.0 (88.8-100.0) | 99.4 (96.9-100.0) | 99.5 (97.4-100.0) |
|  | PC | 100.0 (89.1-100.0) | 99.4 (96.9-100.0) | 97.0 (84.2-99.9) | 100.0 (97.9-100.0) | 99.5 (97.4-100.0) |

### Assessment of utility

MPAs provide results for detection of SARS-CoV-2, influenza, and RSV in a single RT-PCR assay compared to five SoC RT-PCR reactions to get the same information, with variable costs, testing burden, and implementation parameters (Table 4). Manufacturer instructions for GX and TP have strictly defined volume of sample/elution and RNA. PC, similar to SoC, allows the use of different sample volumes according to extraction kit. GX and TP are designed for manufacturer-specific instruments. SoC and PC can be utilized on any instrument providing more than two and four optical channels respectively. Run time of 90min per run for 94 samples with TP and PC is similar to SoC but simultaneously provide results for all targets. GX integrates the process from extraction to result, but only for one sample per run. TP required specific training to use the QuantStudio 5 and CE-IVD software for interpretation, while SoC and PC were interpreted on current laboratory software.

**TABLE 4:** Comparison of multiplex assays with regards to test specifications, costs and accomplishment

|  | Standard assay | TaqPath | Powerchek | Xpert Xpress |
| --- | --- | --- | --- | --- |
| Manufacturer | - | Thermo Fisher Scientific | Kogene Biotech | Cepheid |
| Pathogen detection | SARS-CoV-2,<br>IAV, IBV<br>RSVA and RSVB | SARS-CoV-2,<br>InfluenzaA/B,<br>RSV | SARS-CoV-2,<br>IAV, IBV<br>RSV | SARS-CoV-2,<br>IAV, IBV<br>RSV |
| Number of PCR reactions | 4 | 1 | 1 | 1 |
| Sample volume | Not specified * | 400 µl | Not specified * | 300 µl |
| Elution volume | Not specified * | 50 µl | Not specified* | *Not applicable* |
| RNA volume | 25µl (5 µl/each<sup>‡</sup>) | 17.5 µl | 5 µl | *Not applicable* |
| Internal control | Not provided | Provided | Provided | Provided in cartridge |
| Step to add IC | Extraction | Extraction | PCR mix | *Not applicable* |
| Number of samples tested on the same assay | 93 | 94 | 94 | 1 |
| Run on time | 90 min | 90 min | 90 min | 36 min |
| Time to result* | 540 minutes** | 145 min** | 145 min** | 40 min |
| Personnel training | Low | High | Low | Low |
| RT-PCR Instrument | Any with > 2 optical channels | Applied Biosystems™ 7500 Fast; QuantStudio™5; QuantStudio™ 7 Flex, 384–well block | Any with 4 optical channels | GenXpert Instrument |
| Interpretation of results with software | RT-PCR Instrument | Pathogen Interpretive | RT-PCR Instrument | GenXpert Instrument |
| Cost reagents per test (US\$) | 20<sup>†‡</sup> | 25<sup>††</sup> | 13.6<sup>††</sup> | 31<sup>‡</sup> |

\*Sample volume according to the extraction kit manufacturer manual for use; \*\* Extraction 35 min, PCR 90 min each, interpretation of results 10 min for SoC, 20 min for MPA Providing that we perform 5 RT-PCR assays (2 PCR for SARS-CoV-2 (E and RdRP genes); 1 for IAV; 1 for IBV; and 1 for RSV) for Standard of care with 5µl for each RT-PCR
† We calculated a cost of 4 \$ per test in Standard PCR assay. ††Shipment and controls in each PCR plate included. ‡ Shipment included
Prices provided by Singapore for TP, Korea for PC and France for GX.

## DISCUSSION

Incorporation of MPAs into routine surveillance of SARS-CoV-2, influenza and RSV is critical to expand pathogen detection while minimizing costs and constrain on human resources within existing capacities/capabilities. A side-by-side comparison of GX, TP and PC using the same large set of viral isolates, including avian influenza, and clinical samples was critical for evaluation, especially for limited resource settings with high probability of AIV spillover.

GX, TP and PC consistently detected all viral lineages of SARS-CoV-2 and influenza; however, GX had slightly higher LoD compared to SoC. Decreased GX testing volume compared to extraction possibly contributed to this discrepancy. Each MPA demonstrated high accuracy to detect all viruses in clinical samples. Overall, median and range of Ct-values obtained with TP and PC were lower than with SoC and GX. Differences in sample volume and lower number of samples tested with GX could affect these values. Discrepancies between assays did occur. One SARS-CoV-2 infection was not detected by PC, and one and four RSV were not detected by TP and GX, respectively. Low viral load (Ct=38-39) by SoC close to LoD and storage issues could impair detectability. Difference in sample testing volume could impact detection with GX. Unfortunately, remaining sample volume did not allow repeated GX testing. Eight samples had a positive result for one target, but were negative with SoC and other MPAs and were considered as false positive results.

Most commercial tests are not specifically designed to identify/distinguish AIV or novel IAV. However, detection of zoonotic AIV infection is paramount, especially in endemic countries such as Cambodia [29], and for pandemic prevention and preparedness globally. GX package insert does assert the test adequately detects AIV [21]; however, PC and TP have no previous data available. This study indicates MPAs can likely identify AIV cases with high accuracy to detect all targets in clinical samples. All variants of SARS-CoV-2 circulating in Cambodia during the collection period were detected.

Previous evaluations of GX reported a high concordance using retrospective clinical samples compared to other Cepheid assays and several MPAs. In the UK [14], Netherlands [30], and Hong Kong [31], GX had 95-99.64% PPA and 100% NPA for targets compared to SoC. No false positive results were observed with GX in this study, but some were previously reported for SARS-CoV-2/RSV co-infections [30,31]. A previous version of PC was evaluated in South Korea with 100% PPA/NPA for SARS-CoV-2, IAV, and IBV and 93.1%/100% for RSV versus comparator [32]. TP has been evaluated using nasopharyngeal specimens with PPA/NPA at 98.2%/100%, 100%/96.5%, and 98.2%/92.8% for SARS-CoV-2, influenza, and RSV, respectively, compared to reference assays [22]. Detection accuracy in the present study of 97%-100% PPA for all targets is similar to these previous findings.

In addition to detection efficiency, MPAs’ utility is critical for routine use in laboratories. Each GX cartridge only tests one sample at-a-time and is more expensive than other MPAs. However, GX provides fastest results with minimal sample handling, an advantage for emergency cases, reduced sample loads, and/or restricted human resources. Moreover, GX does not require extensive expertise in techniques or interpretation. TP and PC minimize volume of RNA required, and significantly reduce instrument occupation time, potentially critical during periods with high testing demand. Result interpretation is provided automatically using specific software for TP and GX, with TP requiring review of amplification curves [22]. PC and SoCs require user interpretation, allowing flexibility but also need expertise to avoid misinterpretation and introduction of potential technical error.

A prospective design was not possible in this study and retrospective investigation was conducted on stored samples, potentially resulting in selection bias and reduced sample quality. This impact was probably limited by selection based on available volume versus specific viral characteristics. Sample volume and/or extracted RNA was too limited to repeat all SoC for all samples, thus some routine results were included from time of reception. However, if any targeted virus was detected with MPA, the same extracted RNA was retested with corresponding SoC. A few samples with low viral loads and limited IBV sample number restricted some further investigations. Finally, determination of LoD by viral copy number requires extensive *in vitro* assessment and electron microscopy, which is not readily available in Cambodia. Future experiments with tittered viral isolates will add to the assessment of LoD.

The reality of overlapping clinical presentations of concurrently circulating viruses, funding and reagent constraints, and limited human resources require integration of MPAs into routine VRI surveillance. Timely diagnosis decreases unnecessary laboratory testing, minimizes use of antibiotics, and maximizes effectiveness of measures to control infection. Appropriate and early antiviral treatment reduces complications, hospitalizations, and mortality [33]. Simultaneous detection of SARS-CoV-2, influenza, and RSV in a single test accelerates time from sampling to diagnosis, and can utilize capacity/capability developed during the COVID-19 pandemic. MPAs also preserve consumables, and streamline human resources to respond to other endemic or emerging pathogens. As result, MPAs have the potential to sustain and even expand surveillance systems, thereby strengthening understanding of seasonal pathogens, availability for vaccine development, and epidemic/pandemic preparedness, prevention, and response.

## Data Availability

All data produced in the present work are contained in the manuscript

## ACKNOWLEDGMENTS

The authors would like to acknowledge the Cambodian Ministry of Health and Cambodian Communicable Disease Control Department, Rapid Response Team members, all of the Provincial Health Departments, and the doctors, nurses, and staff involved in the COVID-19 response in Cambodia. We thank all of patients who supplied samples for this evaluation. We also thank the technicians and staff at Institut Pasteur du Cambodge in the Virology and Epidemiology/Public Health Units involved in this work, especially all of the tirelessly dedicated COVID and Influenza teams.

## CONFLICT OF INTEREST

This work was supported by Thermo Fisher Scientific, who loaned the QuantStudio 5™ RT-PCR, 96 well, 0.2 mL instrument (Applied Biosystems) and laptop to IPC for the purpose of the study, provided TaqPath™ COVID-19, FluA/B, RSV Combo Kits, and co-authors included were involved in the study design, analysis and interpretation of TaqPath results and reviewed the report.

## FUNDING SOURCE

This work was supported by Thermo Fisher Scientific, who loaned the QuantStudio 5™ RT-PCR, 96 well, 0.2 mL instrument (Applied Biosystems) and laptop to IPC for the purpose of the study, provided TaqPath™ COVID-19, FluA/B, RSV Combo Kits, and co-authors included were involved in the study design, analysis and interpretation of TaqPath results and reviewed the report. French Agency for Development funded the ECOMORE II project and a COVID-19 top-up (project No. CZZ 2146 01A), providing payed salary to NB for a senior medical virologist position under a temporary contract with the Institut Pasteur du Cambodge. H. A. is supported by the German Centre for International Migration and Development (CIM). Influenza, COVID-19, and RSV work at IPC and E.A.K are supported, in part, by the World Health Organization and the Food and Agriculture Association of the United Nations. COVID-19 work is supported, in part, E.A.K is funded, in part, by the Pasteur International Center for Research on Emerging Infectious Diseases (PICREID) National Institutes of Health, Department of Health and Human Services -funded project (*1U01AI151758*-01).

## ETHICAL APPROVAL STATEMENT

This study was approved by the Cambodian National Ethics Committee for Health Research (N°050 NECHR, 2022). Since samples were obtained as part of the national influenza surveillance system and as part of outbreak response for SARS-CoV-2, requirement for informed consent was waived for their use in the study. All samples were de-identified and the database contained no patient information.

## Notes

### Competing Interest Statement

This work was supported by Thermo Fisher Scientific, who loaned the QuantStudio 5TM RT-PCR, 96 well, 0.2 mL instrument (Applied Biosystems) and laptop to IPC for the purpose of the study, provided TaqPathTM COVID-19, FluA/B, RSV Combo Kits, and co-authors included were involved in the study design, analysis and interpretation of TaqPath results and reviewed the report.

### Funding Statement

This work was supported by Thermo Fisher Scientific, who loaned the QuantStudio 5TM RT-PCR, 96 well, 0.2 mL instrument (Applied Biosystems) and laptop to IPC for the purpose of the study, provided TaqPathTM COVID-19, FluA/B, RSV Combo Kits, and co-authors included were involved in the study design, analysis and interpretation of TaqPath results and reviewed the report. French Agency for Development funded the ECOMORE II project and a COVID-19 top-up (project No. CZZ 2146 01A), providing payed salary to NB for a senior medical virologist position under a temporary contract with the Institut Pasteur du Cambodge. H. A. is supported by the German Centre for International Migration and Development (CIM). Influenza, COVID-19, and RSV work at IPC and E.A.K are supported, in part, by the World Health Organization and the Food and Agriculture Association of the United Nations. COVID-19 work is supported, in part, E.A.K is funded, in part, by the Pasteur International Center for Research on Emerging Infectious Diseases (PICREID) National Institutes of Health, Department of Health and Human Services -funded project (1U01AI151758-01).

### Author Declarations

This study was approved by the Cambodian National Ethics Committee for Health Research (No050 NECHR, 2022). Since samples were obtained as part of the national influenza surveillance system and as part of outbreak response for SARS-CoV-2, requirement for informed consent was waived for their use in the study. All samples were de-identified and the database contained no patient information.

