## Supplemental Files for "ONE ASSAY TO TEST THEM ALL: COMPARING MULTIPLEX ASSAYS FOR EXPANSION OF RESPIRATORY VIRUS SURVEILLANCE"

**SUPLEMENTARY TABLE 1**:

| Virus | n | Co-infection | Variant/subtype |
|---|---|---|---|
| SARS-CoV-2 obtained between February 2020 to June 2022 | 56 | 0 | Wuhan n=10;<br>Alpha n=12;<br>Delta n=12;<br>Omicron n= 22<br>(sub-variants BA.1 n=7; BA.2 n=6; BA.2.12.1 n=3; BA.4 n=1; BA.5 n=5) |
| IAV<br>from 2016 to 2019 | 84 | RSV n = 4<br>(RSVA n= 3;<br>RSVB n = 1) | H1N1 n=82<br>H3N2 n=2 |
| IBV<br>from 2016 to 2019 | 5 | 0 | Victoria n=4<br>Yamagata n=1 |
| RSV<br>from 2016 to 2022 | 28 | SARS-CoV-2 n= 2 | RSVA n=29<br>RSVB n=3 |
| None (negative)<br>from 2019 to 2022 | 126 | 0 | - |

**Legend:** SARS-CoV-2: severe acute respiratory syndrome coronavirus 2, IAV: Influenza A; IBV: Influenza B; RSVA/B: respiratory syncytial virus

**SUPPLEMENTAL TABLE 2**: Detection of samples per target for each assay:

| | | SOC | | | FAO | | | TP† | | | PC | | | GX | | |
|---|---|---|---|---|---|---|---|---|---|---|---|---|---|---|---|---|
| | | + | - | total | + | - | total | + | - | total | + | - | total | + | - | total |
| **SARS-CoV-2** | + | 59 | 0 | 59 | - | - | - | 57 | 2 | 59 | 56 | 3 | 59 | 57 | 2 | 59 |
| | - | 0 | 180 | 180 | - | - | - | 0 | 180 | 180 | 1 | 179 | 180 | 0 | 165 | 165 |
| | total | 59 | 180 | 239 | - | - | - | 57 | 182 | 239 | 57 | 182 | 239 | 57 | 167 | 224 |
| **IAV** | + | 84 | 0 | 84 | 48 | 36 | 84 | - | - | - | 84 | 0 | 84 | 53* | 0 | 53 |
| | - | 0 | 179 | 179 | 0 | 0 | 0 | - | - | - | 1 | 178 | 179 | 0 | 179 | 179 |
| | total | 84 | 179 | 263 | 48 | 36 | 84 | - | - | - | 85 | 178 | 263 | 53 | 179 | 232 |
| **IBV** | + | 5 | 0 | 5 | - | - | - | - | - | - | 5 | 0 | 5 | 5 | 0 | 5 |
| | - | 0 | 222 | 222 | - | - | - | - | - | - | 3 | 219 | 222 | 0 | 203 | 203 |
| | total | 5 | 222 | 227 | - | - | - | - | - | - | 8 | 219 | 227 | 5 | 203 | 208 |
| **Influenza†** | + | 89 | 0 | 89 | - | - | - | 89 | 0 | 89 | - | - | - | - | - | - |
| | - | 0 | 174 | 174 | - | - | - | 0 | 174 | 174 | - | - | - | - | - | - |
| | total | 89 | 174 | 263 | - | - | - | 89 | 174 | 236 | - | - | - | - | - | - |
| **RSV** | + | 32 | 0 | 32 | - | - | - | 31 | 1 | 32 | 32 | 0 | 32 | 28 | 4 | 32 |
| | - | 0 | 178 | 178 | - | - | - | 0 | 178 | 178 | 1 | 177 | 178 | 0 | 163 | 163 |
| | total | 32 | 178 | 210 | - | - | - | 31 | 179 | 210 | 33 | 177 | 210 | 28 | 167 | 195 |

**Legend:** SARS-CoV-2: severe acute respiratory syndrome coronavirus 2, IAV: Influenza A; IBV: Influenza B; RSVA/B: respiratory syncytial virus; SOC: standard or care (Berlin for Sars-CoV-2; CDC for IAV and IBV, Duplex VIDRL for RSV); FOA: Food and Agriculture Organization of United Nations recommended primers and probes developed by the Australian Center for Disease Prevention for the detection of M gene from avian influenza; GX: Xpert® Xpress SARS-CoV-2, Flu, RSV Kit; TP: TaqPath™ COVID-19, FluA/B, RSV Combo Kit; PC: PowerChek™ SARS-CoV-2, Influenza A&B, RSV Multiplex Real-time PCR Kit II. * For 31 specimens, the volume of sample available did not allow us to perform the GX test. † TaqPath does not provide separate results for IAV and IBV, as targets are combined in the same optical detection channel. Therefore, IAV and IBV samples were analyzed conjointly for comparison with SoC.

**SUPPLEMENTAL TABLE 3:** Discordant results among clinical specimens

| Sample ID | Discordance type | SARS-CoV-2 | | | | Influenza | | | | RSV | | | |
|---|---|---|---|---|---|---|---|---|---|---|---|---|---|
| | | SOC | GX | TP | PC | SOC | GX | TP† | PC | SOC | GX* | TP* | PC* |
| 4 | **SARS-CoV-2** | nd | nd | nd | **+ (34)** | nd | nd | nd | nd | + (21) | + (25) | + (15) | + (13) |
| 5 | | + (38) | **nd** | **nd** | **nd** | **nd** | nd | nd | nd | + (22) | + (29) | + (16) | + (13) |
| 13 | | + (39) | + (37) | + (33) | **nd** | **nd** | nd | nd | nd | + (33) | + (33) | + (28) | + (25) |
| 29 | **Influenza** | nd | nd | nd | nd | nd<br>IBV (30) | nd<br>IBV (26) | Inf (32) | **IAV (33)**<br>IBV (26) | nd | nd | nd | nd |
| 40 | | + (33/37) | + (35) | + (32) | + (28) | nd | nd | **Inf (13)** | nd | nd | nd | nd | nd |
| 126 | | nd | nd | nd | nd | IAV (20)<br>nd | IAV (20/25)<br>nd | Inf (11) | IAV (14)<br>**IBV (31)** | nd | nd | nd | nd |
| 162 | | nd | nd | nd | nd | IAV (21)<br>nd | IAV (22/28)<br>nd | Inf (12) | IAV (17)<br>**IBV (31)** | nd | nd | nd | nd |
| 221 | | nd | nd | nd | nd | nd | nd | nd | nd<br>**IBV (34)** | nd | nd | nd | nd |
| 8 | **RSV** | nd | nd | nd | nd | nd | nd | nd | nd | RSVA (30) | **nd** | + (25) | + (22) |
| 17 | | nd | nd | nd | nd | nd | nd | nd | nd | RSVA (26) | **nd** | + (23) | + (17) |
| 27 | | nd | nd | nd | nd | nd | nd | nd | nd | RSVA (30) | **nd** | + (24) | + (20) |
| 73 | | + (35/36) | + (32) | + (24) | + (26) | nd | nd | nd | nd | nd | nd | **+ (35)** | nd |
| 127 | | nd | nd | nd | nd | IAV (28) | IAV (27/29) | Inf (13) | IAV (22) | nd | nd | nd | **+ (33)** |
| 146 | | nd | nd | nd | nd | IAV (24) | IAV (25/28) | Inf (18) | IAV (19) | RSVA (39) | **nd** | **nd** | + (38) |

**Legend:** SoC: standard or care (Berlin for Sars-CoV-2; CDC for IAV and IBV, Duplex VIDRL for RSV); GX: Xpert® Xpress SARS-CoV-2, Flu, RSV Kit; TP: TaqPath™ COVID-19, FluA/B, RSV Combo Kit; PC: PowerChek™ SARS-CoV-2, Influenza A&B, RSV Multiplex Real-time PCR Kit II; SARS-CoV-2: severe acute respiratory syndrome coronavirus 2, Flu: influenza, IAV: Influenza A; IBV: Influenza B; RSV/RSVA: respiratory syncytial virus; nd: not detected; † TP uses the same optical channel for the detection of IAV and IBV and therefore provides a combined result for influenza.* GX, TP and PC use the same optical channel for the detection of RSVA and RSVB and then provide a combined result for RSV.
